# Estimating Under-diagnosis of Patients in Chronically-ill Populations

**DOI:** 10.1101/2022.01.25.22269859

**Authors:** Andrew Stocking, Ian Duncan, Nhan Huynh

**Affiliations:** Principle Business Enterprises, Inc.; Department of Statistics and Applied Probability, University of California Santa Barbara; Santa Barbara Actuaries Inc.

## Abstract

Diagnosis coding in administrative data is known to be inconsistent and incomplete, introducing inaccurate assessment of patients’ health outcomes. The under-diagnosis of members with a target chronic condition reduces the correlation of that chronic condition with associated events. Yet, only a few studies have evaluated the extent of under-reporting of chronic conditions in administrative data. In this study, we developed a novel framework to identify latent members, or those who have not yet been identified with a target chronic condition through claims-based diagnosis but are likely to have some degree of the condition. We applied our innovative approach to estimate the prevalence of a chronic-related event, based on the population of observed and latent members. We provided a detailed illustration that treats incontinence as our target chronic condition while examining four types of incontinence-related events: urinary-tract infections (UTIs), slips and falls, dermatitis, and behavioral disruptions. All analysis relied on the 5% Medicare sample for a continuously enrolled cohort between 2014-2018. Using our novel approach, we were able to increase our identification of incontinence from the 11.2% diagnosed in 2018 to an estimated prevalence in 2018 of 34.7% among fee-for-service Medicare beneficiaries over the age of 65. Similarly, our estimation of UTIs associated with those with incontinence increased from 38% to 68%, from 20% to 41% for IAD, 22% to 54% for slips and falls, and from 26% to 57% for behavior disruptions.

## BACKGROUND

Many studies of chronically-ill populations use administrative data (claims) rather than clinical records because of the availability of large, curated datasets (and corresponding unavailability of large clinical datasets). The issue with claims-based identification of incidence and prevalence of a chronic condition is that it depends on the claim-based coding of the condition during the period of observation. Coding practices vary, as do patient compliance and provider access, resulting in incomplete annual coding of conditions. Some reimbursement systems, such as Medicare Advantage’s risk-adjusted payments that depend on patient risk profiles as identified through claims-based diagnoses provide direct incentives to providers for complete and accurate coding and disincentives for inaccurate or exaggerated coding. This is not true, for example, of the Medicare fee-for-service population, as demonstrated amply in the literature [1-5]. We propose a method to identify more accurately the true prevalence of patients with a chronic condition in a claims dataset by identifying those we call “latent” members, members who likely have the condition who are not currently identified through claims but who will be identified in the future (or have been previously). We demonstrate the method with an application to patients with incontinence.

Incontinence is likely to be a more extreme case of under-reporting for several reasons. First, for many people, incontinence often begins light and becomes more severe over time. For example, stress incontinence occurs because of a weak pelvic floor that can cause bladder to leak during exercise, coughing, sneezing, laughing, or any body movement that puts pressure on the bladder. This type of incontinence can be under-reported because it may be embarrassing to discuss, is not observed during a primary care visit, or may be considered a natural part of the aging process [6, 7]. Second, there is no HCC group for incontinence in the CMS reimbursement model, further reducing the incentive to report it. That is, even within the MA setting there is no reimbursement benefit to an incontinence diagnosis which reduces the likelihood that primary care providers or other providers have developed methods to accurately identify incontinence.

### Estimation of under-diagnosis and under-reporting of chronic conditions

The issue of under-coding and over-coding has been studied intensively in Medicare. Medicare fee-for-service physicians are reimbursed on the basis of resource value units for each CPT code that they submit.^4^ Although a diagnosis is required for a claim to be reimbursed, only a single diagnosis is required for physician services. Furthermore reimbursement is based on *units of work* as embodied in the CPT code rather than the patient complexity (comorbidity conditions). A physician can see a complex polychronic patient 100 times a year and provide a diagnosis of diabetes for every visit and be paid the same amount (for the same CPT code), even though the patient has six other comorbidities that contribute to their health. The patient’s comorbid heart condition, hypertension and other conditions are not required for reimbursement and will not be recorded. Fee-for-service reimbursement, therefore, does not provide an incentive for recording patient diagnoses and complexity. The fee-for-service system rewards the “inputs” of care regardless of patient complexity or clinical outcome. Medicare Advantage (MA) plans, conversely, are reimbursed on the basis of a risk-adjusted premium. Risk adjustment takes account of a member’s conditions, as long as they are recorded in claims. MA plans are likely to record a more complete range of diagnosis codes than FFS Medicare (See for example [1-5, 8-12]).

Applying to the model a broader range of recorded diagnoses for an individual with otherwise equivalent risk factors (including health status) is likely to result in a higher risk score for the individual with the wider range of diagnoses.^5^

As discussed above, Congress has recognized that differences in coding (“coding intensity”) will potentially result in non-equivalence between a plan’s members and members with the same health profile in FFS Medicare; and, for this reason, requires a coding pattern adjustment in MA. Differences exist not just in coding intensity between fee-for-service and MA systems but also between different providers in the MA system. Even with more complete coding, patients do not always seek treatment for their conditions on an annual basis (the typical time-period of a population-based study). This leads to under-reporting of chronic conditions, even when coding is complete. An example from the early 2000s (Duncan [12]; see Kronick [11] for a more recent example) illustrates this issue. Table 1 compares the patients identified in adjacent years using three algorithms: Narrow (medical claims with 2 or more encounters for the same diagnosis in the year), Broad (medical claims with 1 or more encounters for the same diagnosis in the year) and Rx (Broad definition + 3 or more prescriptions for an appropriate medication). As the table shows, using a typical (narrow) definition only three-quarters of patients re-identify in a second year; even with a very broad definition all patients are not re-identified.

**Table 1:** Identification of Chronic Patients in Adjacent Years

| Prevalence of Claims-based identification in a Chronic Population |  |  |  |  |  |
| --- | --- | --- | --- | --- | --- |
| Year 1 |  | Narrow | + Broad | + R<sub>X</sub> | TOTAL |
| Year 2 | Narrow | 75.9% |  |  |  |
|  | + Broad |  | 85.5% |  |  |
|  | + R<sub>X</sub> |  |  | 92.9% |  |
|  | Not Identified | 24.1% | 14.5% | 7.4% |  |
|  | TOTAL | 100.0% | 100.0% | 100.0% | 100.0% |

A second source of identification of under-reporting of chronic patients is the comparison of claims-based identification with national statistics. This type of comparison should be approached with caution because national statistics include patients from a variety of payers (including Medicare, Medicaid, Commercially-insured (employer) populations and the uninsured). As an example, prevalence of urinary incontinence (UI) in a sample of 1,243,173 Medicare fee-for-service lives over age 65 amounted to 11.2%, compared with national statistics of 60% for women and 35% for men [13].

## METHODOLOGY

### Estimating prevalence of chronic conditions

Recognizing that true prevalence of a chronic condition through claims may be underestimated, we propose a method for estimating prevalence. We do so using an historical cohort of patients. We identify a longitudinal cohort of continuously enrolled members of the population in Δ years where *Ct* represents the number of members in the baseline year *t*. Note that our setup of using a continuous cohort helps simplify the estimation approach but under-estimation is still possible (see Discussion section for further explanations). With this setup, the size of our cohort remains constant over Δ years of the study: *C*^*t*^ = *C* ^*t* +*i*^ for *i* = 0,…, Δ −1. Let 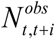 be the observed number of members who are newly-diagnosed with the target condition in year *t* + *i* (*i* = 0,…, Δ −1) and experienced the condition-related events in the baseline year *t*. We illustrate our setup in Figure 1.

**Figure 1:**
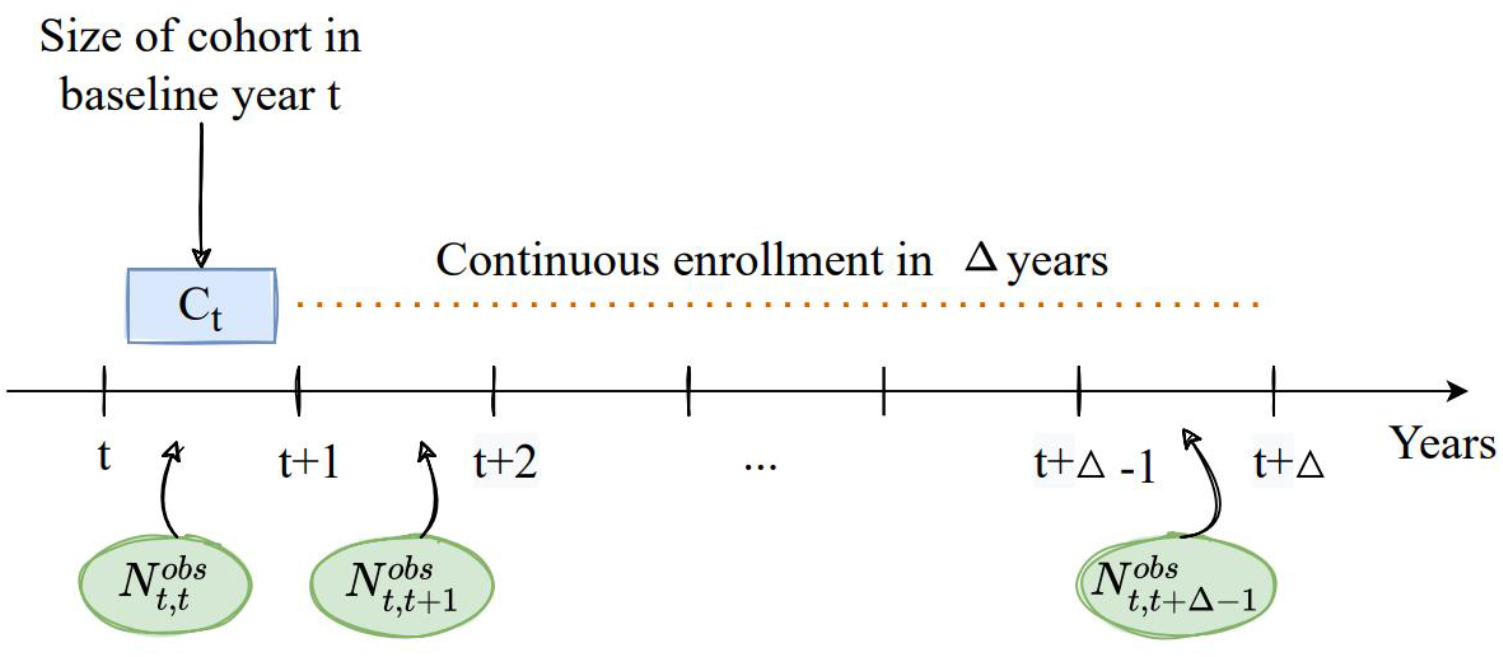
Timeline of members diagnosed with the target chronic condition during the study

**Figure 2:**
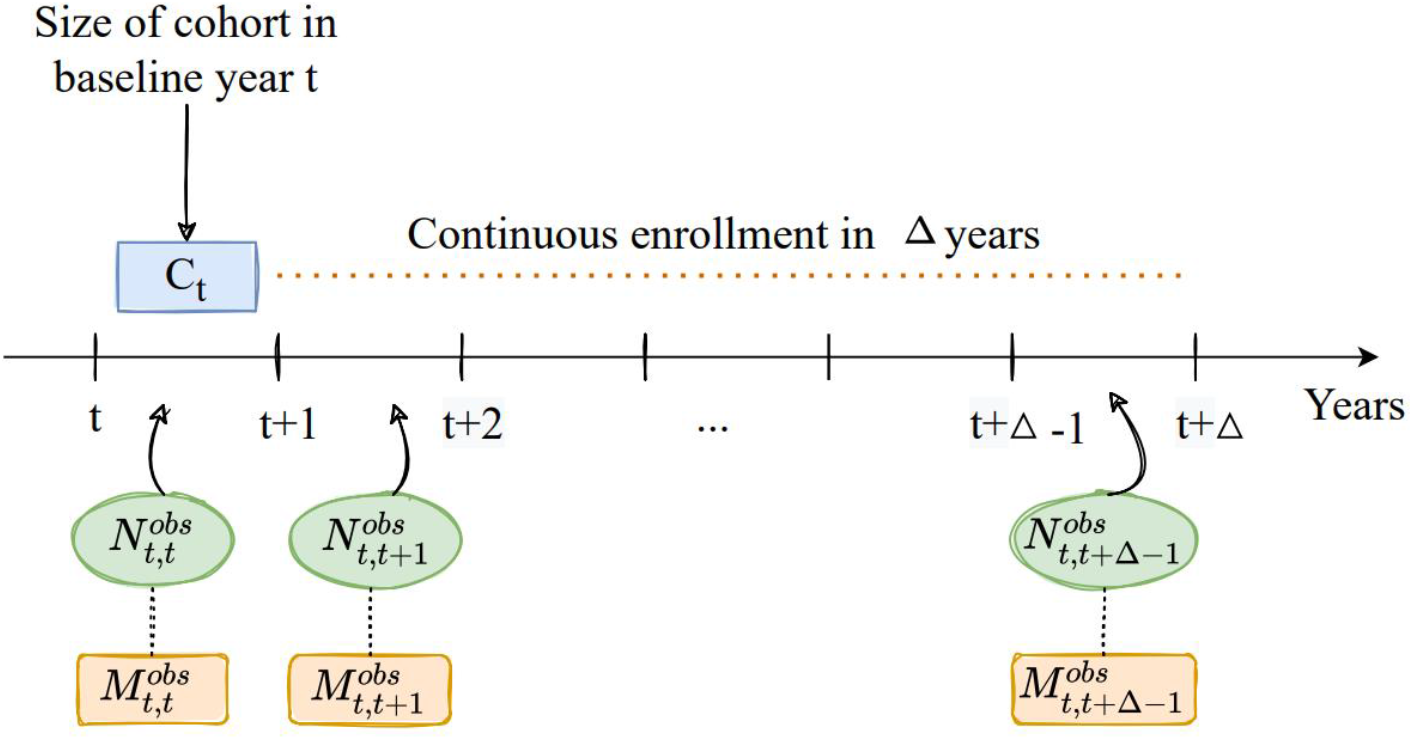
Association between chronic-related events and members diagnosed with the condition in the study

Let *N*_*t*_ be the *true* number of members who should have been diagnosed with the target condition in the baseline year *t*. We can express *N*_*t*_ as follows:

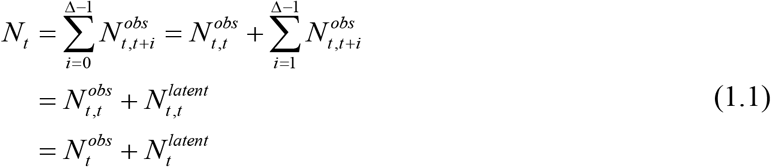

In Eq. (1.1), *N*_*t*_ can be decomposed into the observed number of members diagnosed with the condition in the baseline year 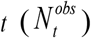 and the latent number of members in the baseline year 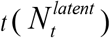. Intuitively, we identify the latent members by adding up all cases of members with the delayed diagnosis of the condition in all future years *t* + *i* (*i* =1,…, Δ −1).

Let 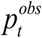 be the observed prevalence of the target condition in the baseline year *t*, then we can express the prevalence in terms of the number of members in the cohort identified with the condition and the size of the cohort, 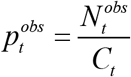. Similarly, we denote *p*_*t*_ as the *true* prevalence of the target condition in year *t* and 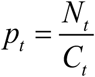. The ratio of the true prevalence to the observed prevalence is:

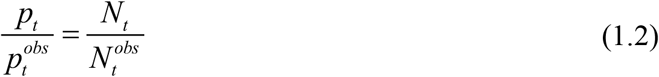

Looking into the future year *k* (*k* > *t*), the relationship between the true and the observed number of members diagnosed with the condition can be expressed as: 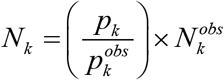. Assuming the prevalence ratio in year *k* remains the same as in the baseline year *t* and the pattern of under-coding in fee-for-service claims persists, we can partially estimate the members identified with the chronic condition in year *k* as:

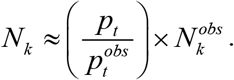

### Effect of under-diagnosis on the reporting of associated events

The under-identification of members with the condition results in corresponding under-attribution of associated events to the chronic condition.^6^ The attribution of events matters for different reporting purposes, particularly for quality reporting and for the types of contracts that rely on quality measures.

#### Appropriately attributing events to identified latent members

Let 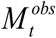 be the observed number of events associated with members identified with the condition in the baseline year *t*. Then, the relationship between 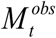 and 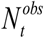 is:

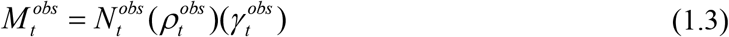

where 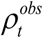 is the frequency of members diagnosed with the condition experiencing the event in the baseline year *t* and 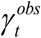 is the conditional average number of event encounters. When the target chronic condition is decomposed into multiple types (e.g. diabetes with three hierarchical groupings in the CMS-HCC risk adjustment models), Eq. (2) can be generalized as follows to incorporate different types of the chronic condition:

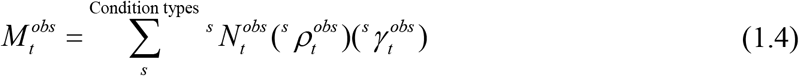

In Eq (1.4) above:

- 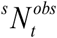 represents the observed number of members with the type-*s* condition and 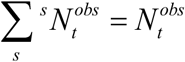
- 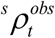 is the frequency of members diagnosed with the condition type *s* experiencing the event
- 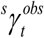 is the conditional average number of event encounters for members with the target condition type *s*.

We denote *M*_*t*_ as the total count of the chronic-related event in the baseline year *t*. Calculating *M*_*t*_ involves two populations: members diagnosed with the condition in the baseline year *t* while experiencing the event in the same year and members with the delayed diagnosis in the future years *t* + *i* (*i* =1,…, Δ −1) while experiencing the events in the baseline year *t* (See Table 2 for illustration). Employing similar setup described in Eq. (1.1), we can express *M*_*t*_ as follows:

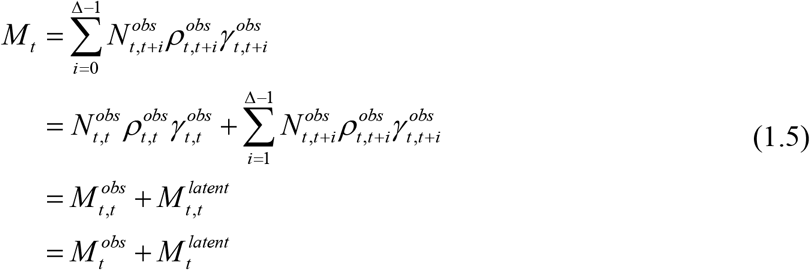

**Table 2:**
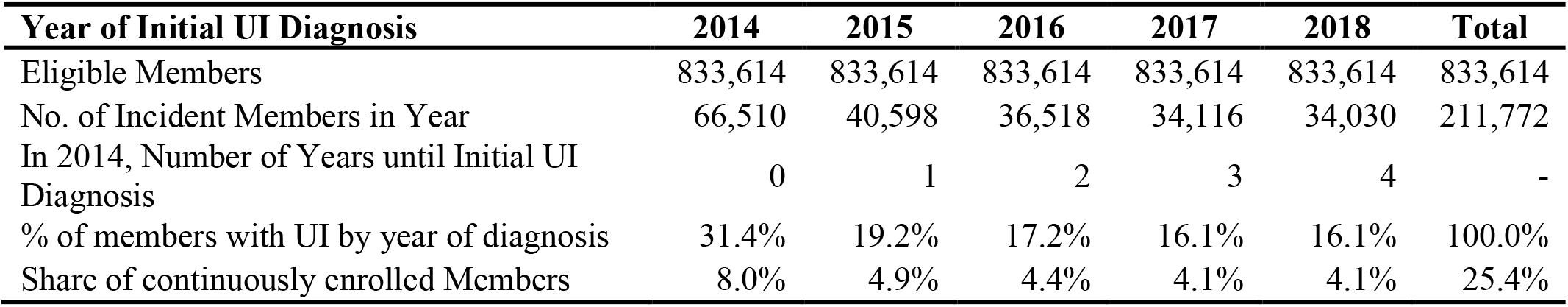
Identification of Incident UI Members by Year

Through the claims-based diagnosis in the baseline year 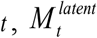 can be thought of as the number of events associated with members without a claims-identified target condition. Thus, under-identification of members with the condition can under-estimate the prevalence of events attributable to the condition population and over-estimate the events in the non-condition population, see Figure 3.

**Figure 3:**
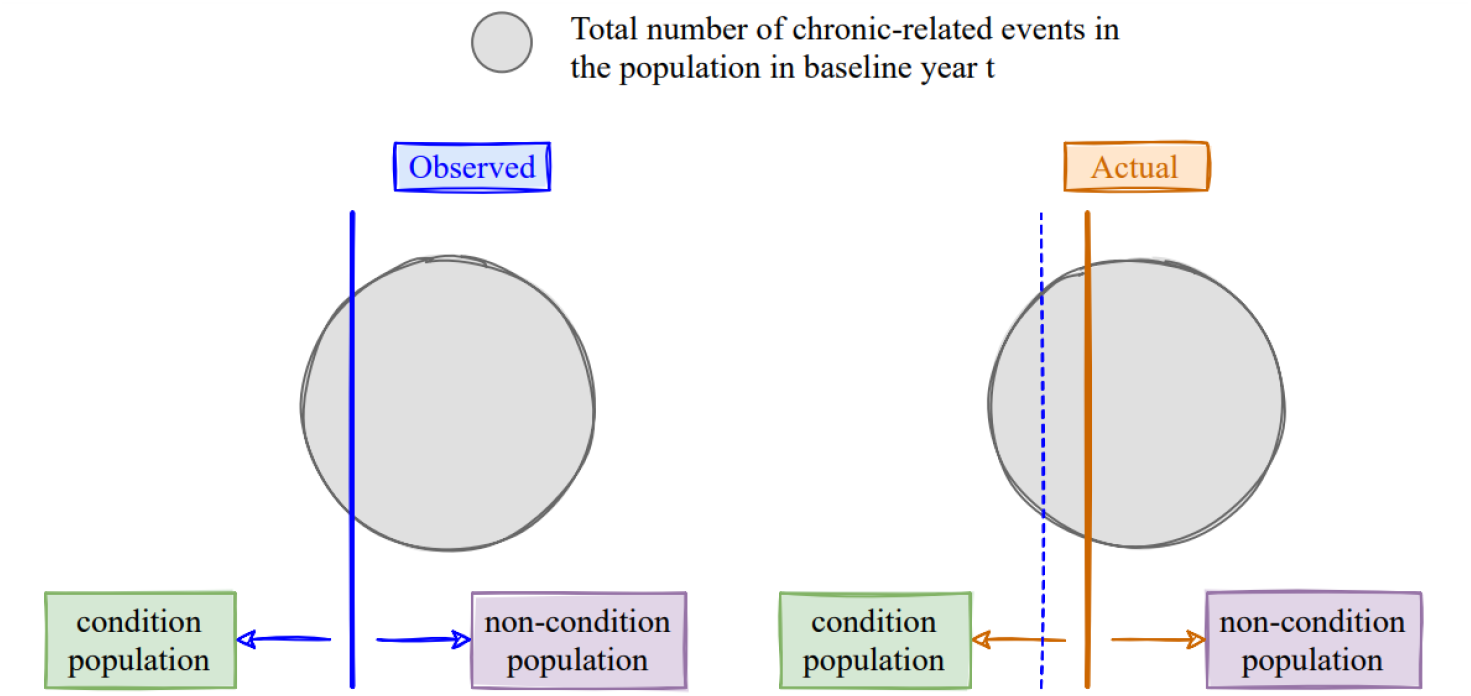
Effect of under-diagnosis on the reporting of associated events

We can infer the relationship between the total and the observed number of chronic-related events in future year *k* (*t* < *k* ≤ Δ − 1) and the baseline year *t* :

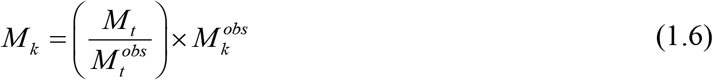

In the above equation, 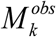 is the observed number of the chronic-related events in the year *k* and *M*_*k*_ is the total number of the chronic-related events in year *k*. Note that 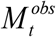 and 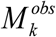 are straightforward to estimate using claims-based diagnosis in year *t* and *k*, respectively; while *M*_*t*_ can be estimated using calculation approach in Eq. (1.5).

## AN APPLICATION TO THE UNDER-DIAGNOSIS OF INCONTINENCE

### Study design

For this study, we evaluated the extent of under-reporting of the prevalence of urinary incontinence (UI) as a result of using a claims-based identification algorithm. We identified urinary incontinence (UI) and fecal incontinence (FI) using ICD-10 codes in the R and N series (see Appendix A). We then estimated the extent of under-reporting of incontinence in a Medicare fee-for-service dataset, limited to members aged 65 and older, by analyzing a continuously enrolled patient cohort. We also required at least 3 months of enrollment in each year per member to reduce duration bias caused by beneficiaries who have minimal exposure to Medicare. All results in our illustration were generated using SAS software version 9.4.

We determined the incidence of four types of incontinence-related events for each cohort: 1) UTIs, 2) Incontinence Associated Dermatitis or Perineal Dermatitis (IAD), 3) Slips, falls and related fractures, and 4) Behavioral disruptions (See Appendix A). Incidence for each event is calculated as the share of members within each cohort who experienced at least one event in the year of the UI/FI diagnosis.

### Estimation of incontinent prevalence

In total there were 833,614 patients continuously enrolled between 2014 and 2018. Thus, the baseline year in this illustration is *t* = 2014, size of our cohort in the baseline year is *C*_2014_ = 833, 614, and duration of the study is Δ = 5 years. We attributed the patient to the year of first identification with an incontinence diagnosis (excluding FI). The resulting patient numbers can be seen in Table 2. In 2014, we have identified 66,510members with incontinent diagnosis and the remaining 767,104 out of 833,614 patients do not have any incontinence claim 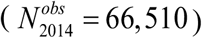. In the following year, of the 767,104 patients 40,598 are identified as incontinent through claims while having incidence of incontinence-related events in 2014 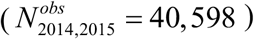. Our hypothesis is that the majority of these patients suffered from some form of incontinence the previous year but had not sought medical help (or generated a claims-based diagnosis) in 2014. By 2018, the last year for which we have data, 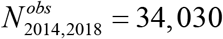.

Applying Eq. (1.1), we can estimate the total number of members who should have identified with incontinence in the baseline year 2014:

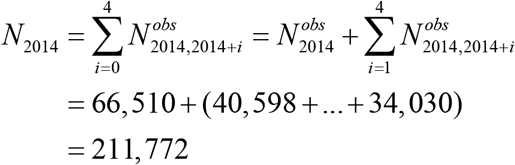

We have identified 145, 262 (= 211, 772 − 66,510) latent members in 2014. The ratio between the true and the observed prevalence of incontinence in 2014 is 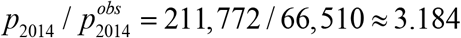. Analyzing 2018 claims data, the number of incident patients identified is 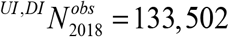 (excluding FI members). Thus, we can estimate the ultimate number of condition members that will emerge within the following four years from 2018 incidence cohort as:

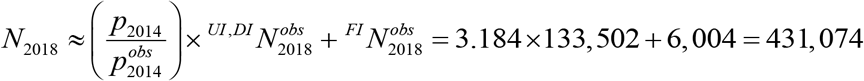

The 2018 latent UI/DI members is 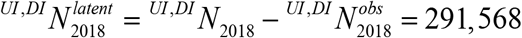 and the prevalence of incontinence is 431, 074 /1, 243,173 ≈ 34.7%.

### Estimation of prevalence of incontinence-related events: UTI, Slips & Falls, Dermatitis, and Behavioral Disruptions

Identification of latent members allows us more accurately to assign the complications of events associated with incontinence to the incontinent and latent incontinent population. Exacerbations that are associated with UI/FI are urinary tract infections, slips and falls, dermatitis and behavior disruptions [14-17]. We reported the frequency of these events among the 2018 incident patients in Table 3. Let us further assume that the distribution of 2018 latent population is the same as the 2014 cohort (Table 2) and the latent members emerge with claims-based diagnoses between 2019-2022 with the same frequency as the 2014 cohort.

**Table 3:** Incontinence-event Frequencies in 2018

| 2018 | Urinary Incontinence | Fecal Incontinence | Dual Incontinence | Total Incontinence | Total Non-Incontinence | Total Events |
| --- | --- | --- | --- | --- | --- | --- |
| Members with the condition | 125,412 | 6,004 | 8,090 | 139,506 | 1,103,667 | - |
| **Events related to Incontinence** |  |  |  |  |  |  |
| UTI | 211,886 | 6,002 | 21,739 | 239,627 | 381,721 | 621,349 |
| IAD | 16,030 | 830 | 1,762 | 18,622 | 73,349 | 91,971 |
| Slips & Falls | 313,600 | 13,506 | 34,827 | 361,933 | 1,267,154 | 1,629,088 |
| Behavior disruptions | 34,064 | 1,224 | 6,528 | 41,817 | 117,693 | 159,510 |

Consider UTI as the incontinence-event of interest. From Table 3, the observed number of UTIs associated with members diagnosed with UI/DI in 2018 is 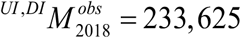. Applying Eq. (1.6), we can approximate the total number of UTIs in 2018 such as:

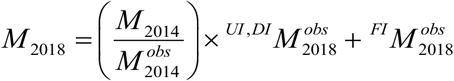

Based on retrospective data analysis in Table 4, we can infer the following:

**Table 4:**
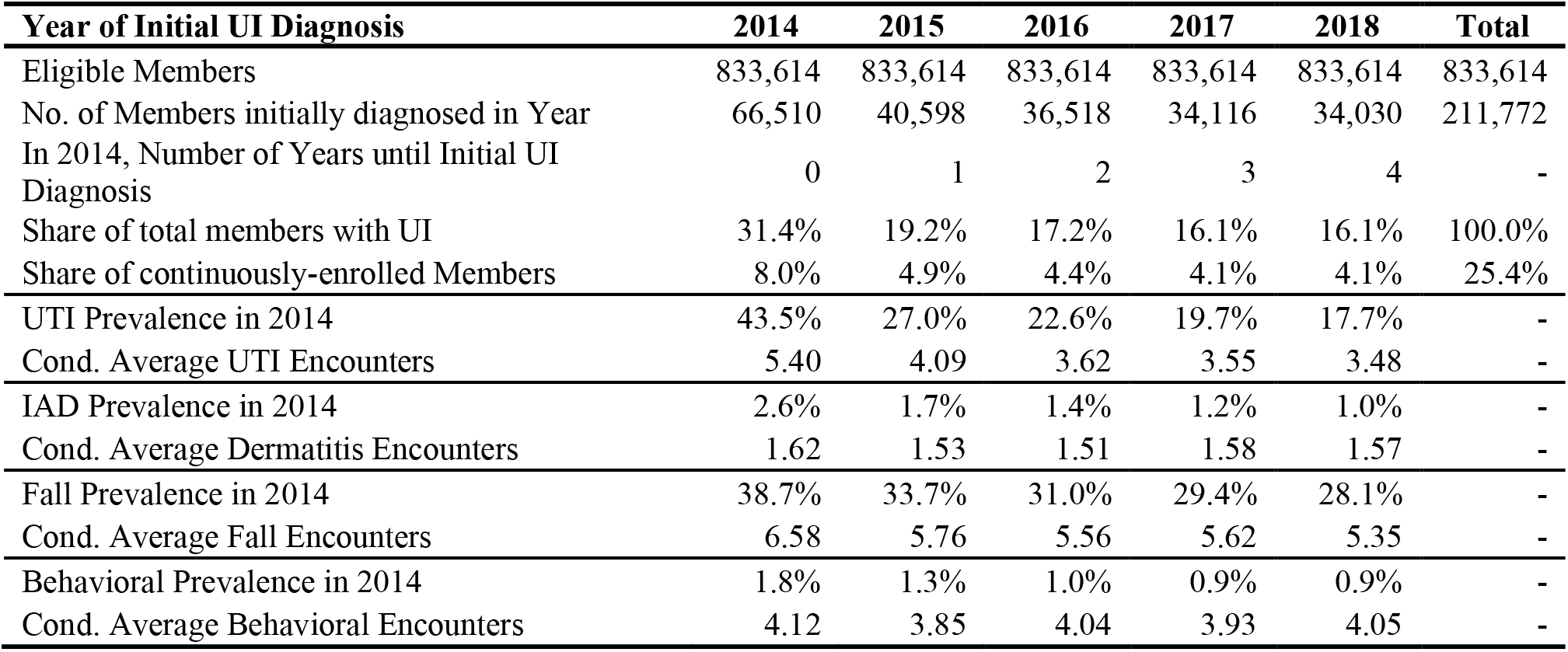
Identification of Incontinence Incidence in the 2014 Cohort

- 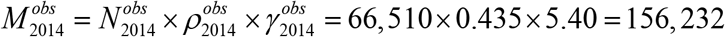
- 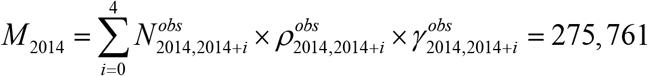

Therefore, *M* = (275,761/156, 232) ×233,625 + 6,002 ≈ 418,367. And 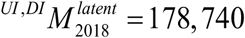 is the number of UTIs associated with the latent UI/DI incontinent. If only UTI events of the observed patients are counted, UTI event prevalence is 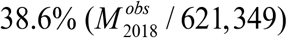; estimating UTI events for the latent incontinence increases UTI prevalence due to the incontinent population to 67.3% (*M*_2018_ / 621,349). Table 5 summarizes the results for other incontinence-related events using our proposed analysis.

**Table 5:** Summary of latent analysis for events related to incontinence

| Events related to incontinence | $M_{2014}^{obs}$ | $M_{2014}$ | $M_{2018}^{obs}$ | $M_{2018}$ | $\left( \frac{M_{2018}^{obs}}{\text{Total events}} \right)$ | $\left( \frac{M_{2018}}{\text{Total events}} \right)$ |
| --- | --- | --- | --- | --- | --- | --- |
| UTI | 156,232 | 275,761 | 239,627 | 422,959 | 38.6% | 68.1% |
| IAD | 2,801 | 5,810 | 18,622 | 38,627 | 20.2% | 41.0% |
| Slips and falls | 169,365 | 418,641 | 361,933 | 894,636 | 22.2% | 53.7% |
| Behavior disruptions | 4,932 | 10,887 | 41,817 | 9,229 | 26.2% | 56.9% |

## DISCUSSION

The novel approach presented here is able to increase the prevalence of a chronic condition without seeking additional datasets for the same members (e.g., clinical or drug databases). The approach rests on several assumptions. First, the approach relies on the hypothesis that the chronic condition is unlikely to develop within a short period of time. That is, the chronic condition is likely to exist one to five years prior to a formal diagnosis. Second, our approach relies on applying a retrospective setting to the future state. Changes in incentives or information that affect the propensity of providers to diagnose a chronic condition will cause the retrospective approach to be a less accurate prediction for the future.

Considering these two assumptions in the context of incontinence highlights the benefit of the approach. First, we assumed that incontinence often begins light and becomes more severe over time, presumably leading to nearly everyone with an incontinent diagnosis in 2018 also being incontinent in the preceding 5 years, albeit possibly to a lesser degree, but without a confirmed claims-based diagnosis. This assumption is supported by the evidence that incontinent in the elderly is a chronic condition that is likely to exist 1 to 5 years prior to a formal diagnosis [18-20].

The results from Table 4 provide further support for this assumption. We find that as patients are further from the year of their first claims-based incontinence diagnosis, the prevalence of the incontinence-related events monotonically decreases, and the conditional average number of events also tends to decrease. In all cases, the incidence and conditional average for incontinence-related events was lowest for those furthest from the year in which they would be diagnosed with incontinence and increased up until the year of diagnosis. This provides some evidence that those diagnosed in later years as incontinent were experiencing the same set of incontinence-related events in 2014 as those who were diagnosed in 2014, only to a lesser extent.

These results provide some insights into a possible additional underlying cause of under-reporting of chronic conditions. They suggest that maybe the clinical diagnosis is incomplete because healthcare providers treat the symptoms without diagnosing the underlying cause. But when the frequency of symptoms increases above a threshold level, providers are more likely to include the related diagnosis in the claim. With respect to incontinence, the data shows a clear trend that as the incidence of each of these incontinence-related events increases, so does the likelihood of a claim-based diagnosis of incontinence.

Second, we assumed that there were no changes in the incentive or information to diagnose incontinence or the incontinence-related events. We analyzed diagnoses of a continuously-enrolled cohort of members between 2014-2018 to estimate the prevalence of latent incontinence in 2018. Our calculations assume the 2018 latent population is the same as the 2014 cohort and the latent members emerge with claims-based diagnoses between 2019-2022 with the same frequency as the 2014 cohort. We believe this to be true for the chronic condition of incontinence; between 2014 and 2018, no new ICD-10-CM codes were created and no new incentives were added to diagnose incontinence. The same is true for UTIs, slips and falls, and behavioral disruptions. However, this was not true for IAD, which had a new ICD-10-CM code developed and added in 2016.^7^ For that reason, the use of this method for predicting the true prevalence of IAD in 2018 may be limited.

### Limitations

Our proposed framework is subjected to several limitations. First, we did not have external data such as clinical or drug records for the same patients to validate and compare our results. However, these datasets are expensive to obtain and for certain chronic conditions, patients are not treated with medication. Second, our approach could still underestimate the size of the latent population due to limited data and the study design. We could not assess the latent members beyond the data period available to us. Our setup of following a continuously enrolled cohort does not allow for the exit of members due to death. For example, if someone were diagnosed after the first year in question, but died prior to the end of the study period, they would be missed in the count of the true prevalence of the chronic condition for the base year. Although considering member exists from the examined cohort could bring us closer to the true prevalence, it requires a more complex estimation approach which is beyond the scope of this paper. Our main objective is to assess the under-diagnosis of the chronic condition and improve the observed prevalence by identifying more members who should have not experienced the delayed diagnosis. Finding the best estimation technique to identify latent members can be a subject of interest for other future studies. Third, the analysis relies on the assumption that the distribution of latent population in a future year is the same as the baseline cohort. This means the underlying populations for both cohorts should be similar. Proper methods must be conducted to validate this assumption.

## CONCLUSION

This is the first study to propose an innovative approach to estimate the true prevalence of the target chronic disease using administrative data. Using incontinence in our illustration, we measured the degree of under-diagnosis of incontinence in the Medicare 5% sample by comparing the observed prevalence against the reported prevalence in the literature. We showcased the implication of under-diagnosis of members with the chronic condition can result in substantial under-count of the associate events. Thus, it is critical for future studies to validate their results when examining prevalence of chronic diseases and comorbidities in administrative data. Understanding the policy or cost implications of these missing diagnoses is a topic of future research and could be applied to many chronic conditions such as obesity and COPD.

## Data Availability

All data produced in the present study are available upon reasonable request to the authors

### Appendix A. ICD-10 Diagnosis Code List for Incontinence and Related Conditions

| Condition | ICD-9 Diagnosis Code | ICD-10 Diagnosis Code |
| --- | --- | --- |
| Urinary Incontinence | 788.30 | R32 Unspecified urinary incontinence |
|  | 788.63 | R39.15 Urge Incontinence |
|  | 788.91 | R39.81 Functional urinary incontinence |
|  | 596.54, 596.59 | N31.9 Neurogenic Bladder |
|  | 596.51 | N32.81 Overactive bladder |
|  | 596.6, 596.7, 596.89 | N32.89 Bladder spasm |
|  | 625.6, 788.32 | N39.3 Stress incontinence (female) (male) |
|  | 788.31 | N39.41 Urge incontinence |
|  | 788.34 | N39.42 Incontinence without sensory awareness |
|  | 788.33 | N39.46 Stress and Urge incontinence |
|  | 788.36 | N39.44 Nocturnal urinary Incontinence |
|  | N/A | N39.492 postural (urinary) incontinence |
|  | 788.39 | N39.498 reflex urinary incontinence |
| Fecal Incontinence | * | R15 Fecal incontinence |
|  | * | R15.0 Incomplete defecation |
|  | * | R15.1 Fecal smearing |
|  | * | R15.2 Fecal urgency |
|  | * | R15.9 Full incontinence of feces |
|  | * | R98.1 Fecal incontinence, nonorganic origin |
|  | * | K59.2 Neurogenic Bowel |
| Urinary Tract Infections | 599.0 | N39.0 Urinary tract infection, site not specified |
|  | 595 | N30 Cystitis |
|  | 597.0, 099.4, 597.8 | N34 Urethritis and urethral syndrome |
| Incontinence Associated Dermatitis or Perineal Dermatitis | 691.0 | L22 Diaper dermatitis |
|  | N/A | L30.9 Dermatitis, unspecified |
|  | N/A | L24.9 Irritant contact dermatitis, unspecified cause |
|  | 112.3 | B37.2 Candidiasis of skin and nail |
|  | 112.1 | B37.3 Candidiasis of vulva and vagina |
| Slips, Falls, and Fractures | N/A | R29.6 Repeated Falls |
|  | V15.88 | Z91.81 history of falling |
|  | E880-E884, E885.9, E886.0, E887-E888, E917.7, E917.8, E929.3 | W00 – W19 Slipping, tripping, stumbling and falls |
|  | 800-826, 830-908, 910-918, 921-929, 950-957, 959, V54.1 | S00 – S99 (Injuries of external causes) |
| Behavioral Disruptions | 294.11 | F02.81 Alzheimer's dementia late onset with behavioral disturbance |
|  | 294.21 | F03.91 Unspecified dementia with behavioral disturbance |
|  | 331.19 | G31.09 Dementia with Parkinsonism plus<br>F02.81 dementia with behavioral disturbance |
|  | 306.8 | F59 Unspecified behavioral syndromes associated with physiological disturbances and physical factor |
|  | 312.9 | F91.9 Conduct disorder |

## Footnotes

4 Resource value units, or resource-based relative value scale is a Marxian labor-value theory concept introduced by William Hsiao in the late 1980s. (Hsiao WC, Braun P, Dunn D, Becker ER. Resource-Based Relative Values: An Overview. *JAMA*. 1988;260(16):2347–2353).

5 The effect of a broader range of recorded diagnoses will depend on the mapping of diagnoses to HCCs, however. Diagnoses that map to the same HCC would not affect the member’s risk score; diagnoses that map to a more severe HCC within the same diagnostic category, or that map to a different HCC would increase the risk score.

6 For hospital admissions this may not be a significant issue given that hospital claims report a range of diagnoses. Reporting by physicians may be affected in the same way as prevalence.

7 https://www.icd10data.com/ICD10CM/Codes/Changes/New_Codes/134?year=2016

